# Effective cataract surgical coverage in China: results from the China National Eye Health Study (CNEHS)

**DOI:** 10.1101/2024.07.03.24309891

**Authors:** Jiaying Li, Kai Cao, Jie Xu, Xiaobin Yu, Shanshan Jin, Qing Zhang, Ailian Hu, Qinghuai Liu, Haidong Zou, Wenyong Huang, Xiaoling Liang, Zongming Song, Bin Sun, Wenjuan Zhuang, Xiyuan Zhou, Zhulin Hu, Zhengzheng Wu, Hong Zhang, Wei He, Minglian Zhang, Zibing Jin, Ningli Wang

**Author notes:** Primary Corresponding Author: Prof. Ningli Wang, Beijing Tongren Eye Center, Beijing Tongren Hospital, Beijing Ophthalmology and Visual Science Key Lab, Beijing Institute of Ophthalmology, Capital Medical University, NO.1 Dongjiaominxiang Street, Dongcheng District, Beijing, China, 100730;. The authors contributed equally and should be recognized as co-first author (Jiaying Li, Kai Cao). The authors contributed equally and should be recognized as co-corresponding author (Zibing Jin, Ningli Wang).

## Abstract

**Purpose:** The 74th World Health Assembly endorsed a global target for 30% increase in effective cataract surgery coverage (eCSC) from 2020 to 2030. The current study was conducted to help monitoring the progress in the next decade in China.

**Design:** A cross-sectional multi-center study, the China National Eye Health Survey (CNEHS), was conducted from 2021 to 2022 and covered 562 communities/villages from 12 representative provinces in China.

**Methods:** This study is based on participants who were at least 50 years of years in the CNEHS. eCSC estimates the proportion of individuals with operated cataract achieving postoperative visual acuity ≥6/18 out of the total population including both operated and operable cataract cases. Both the CSC and eCSC were adjusted for age and sex.

**Results:** A total of 45,051 participants were included in the analysis. The standardized incidence of vision impairment and blindness (best-corrected visual acuity in the better eye <6/18) was 4.4% (95% CI, 4.2%-4.6%). Cataracts accounted for 52.7% of these cases. The weighted rates of CSC and eCSC were 57.2% (95% CI, 47.6%-66.8%) and 38.7% (95% CI, 31.7%-45.6%), respectively. Contributing factors to the quality gap(difference between CSC and eCSC) included concurrent ocular diseases (62.2%) and complications relevant to surgery. CSC increased by 38.9% and eCSC by 26.1% over the decade. Both CSC and eCSC varied substantially across the 12 provinces (range: 41.2%-87.3% for CSC; 16.7%-71.0% for eCSC). Pearson correlation analysis indicated that regional disparities were primarily explained by the level of medical resource allocation, particularly the number of physicians or ophthalmologists per unit population (r=0.6, p=0.03).

**Conclusion:** During the 2021-2022 period, CSC and eCSC rates in China were 57.2% and 38.7%, respectively. Regional disparity in cataract coverage were observed and correlated with the level of medical resources allocation.

**WHAT IS ALREADY KNOWN ON THIS TOPIC:** The latest national epidemiological data on cataract surgical coverage in China dates back to 2014 and contains limited information.

**WHAT THIS STUDY ADDS:** Between 2021 and 2022, the rates of CSC and eCSC in China were 57.2% and 38.7% at the 6/18 threshold, respectively. Comparing this with historical data reveals a significant improvement, with eCSC increasing by 26.1% over the past decade. However, there remains a notable quality gap, largely due to concurrent ocular diseases (62.2%), which should not be included in the eCSC measure. Additionally, substantial regional disparities were identified, with eCSC rates varying from 16.7% to 71.0% across the 12 surveyed provinces/municipalities. These disparities are primarily linked to the distribution of medical resources per capita rather than economic status.

**HOW THIS STUDY MIGHT AFFECT RESEARCH, PRACTICE, OR POLICY:** Despite significant improvements over the past decade, CSC and eCSC rates in China remain lower than those reported in most other countries. To address this, the government is urged to allocate more medical resources to less developed regions. Furthermore, the definition of eCSC should be revisited to exclude biases from concurrent ocular diseases.

## Introduction

Cataract is the leading cause of reversible blindness worldwide, particularly in low- and middle-income countries due to limited cataract surgical coverage (CSC). Effective cataract surgical coverage (eCSC) includes availability of surgical services but emphasizes the quality and outcomes, and therefore represents an important measure.^1, 2^

The national estimate of CSC using the 6/60 threshold in China was 34.5% in 2006,^3^ and increased to 62.6% in 2014.^4^ Such a rapid improvement could be primarily attributed to advances in healthcare insurance coverage and free cataract surgery projects from both government and non-government organizations. Recently, guided by the 13th Five-Year Plan, efforts have increased to expand surgery coverage and develop ophthalmologic infrastructure, enabling over 90% of rural hospitals to perform cataract surgery. Since the 74th World Health Assembly established a global target of increasing eCSC by 30% before 2030,^5^ it is important to establish baseline number and keep monitor coverage in new decade to ensure effectiveness.

As the first report of China National Eye Health Survey (CNEHS) study, the study aimed to provide a national-level estimate of eCSC, CSC in China, relative quality gap (difference between CSC and eCSC) and regional disparity.

## Methods

### Study design and participants

The CNEHS study is a nation-wide cross-sectional study conducted during a period from May 2021 to September, 2022. The study was approved by the institutional review board of Beijing Tongren Hospital, and conducted following the followed the Declaration of Helsinki. All participants provided written informed consent. Writing of this manuscript complied with the STROBE statement.^6^

A multi-stage cluster sampling technique was utilized to communities/villages from the following 12 provinces/municipalities that represent different geographical areas and levels of socio-economic development (gross domestic production in US dollars from 50,883 to 190,126): Beijing, Hebei, Jiangsu and Guangdong in the East coast, Henan and Shanxi in the inland Middle region, Heilongjiang and Liaoning in the Northeast region, Chongqing, Ningxia, Yunnan and Sichuan in the West region. A total of 562 communities/villages and 89,137 participants were included. A listing of the site coordinators is provided in a eTable1. The current study included 43,045 participants who were 50 years of age or older at the time of the study entry. The sample size requirement was estimated using the following equation:

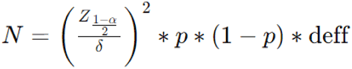

Where:

• N is the sample size;

• δ is the allowable error;

• α is the test level (set at 0.05);

• p is the rate of the primary outcome;

• deff, the design effect, was set at 1.5.

Sample size requirement was calculated separately for the 2 primary outcomes (visual acuity and refractive error), and the higher estimate (62,317) was used. Expecting non-response rate at 20 %,^7–10^ the sample size requirement was 77,896 for total inclusion(aged 18 and above). The response rate of current study is 76.2% and the number of participants included is 89,137.

Quality control in this study was implemented on multiple levels: 1) Pre-survey Training: Uniform training sessions were conducted prior to the survey, covering the operational procedures for various ophthalmologic examinations and questionnaire administration. 2)Electronic Data Collection System: Data collection was conducted using a bespoke electronic data collection system designed specifically for this project. This paperless approach not only saved time and enhanced efficiency but also reduced the likelihood of data entry errors through the system’s logic check capabilities. 3) Automated Data Entry for Specific Measurements: Intraocular pressure and autorefractor data were not manually entered. Instead, a custom Bluetooth module developed for this project automatically captured and stored the measurement results from the devices directly into the electronic data collection system upon completion of each test. 4) Pilot Study: Before the formal survey, a pre-survey was conducted by the project team.

A comprehensive listing of households, including names of residents aged 18 and older who had resided in the area for more than six months, was compiled from village registers. This effort was facilitated by community committees, village committees, and village doctors, who collectively formed the enumeration teams.

### Procedures

Ophthalmic examination was conducted by trained nurses or ophthalmologists, and included visual acuity, refraction, intraocular pressure, slit lamp examination and fundus recordings. Results of the examination were recorded using a standardized form. Presenting distance visual acuity (PVA), with spectacles if the participant had them at presentation, was measured using a retroilluminated logarithm of the minimum angle of resolution (logMAR) “illiterate E” chart. Subjective refraction was used to achieve best-corrected visual acuity (BCVA). Corneal abnormality, cataract or severe amblyopia were determined by ophthalmologist on site and fundus images were sent to a reading center at Tongren Hospital for diagnosis. The principal cause of blindness or vision impairment(VI) for each eye was classified into one of the following 14 categories: refractive error, amblyopia, cataract or posterior capsule opacification(PCO), corneal opacity or scar, glaucoma, other optic atrophy, diabetic retinopathy, age-related macular degeneration, myopic retinopathy, uveitis, ocular trauma, other retinal or choroidal changes, other causes, and undetermined. When multiple causes appeared to be involved, the principal cause of visual impairment favored the most treatable cause.

Blindness was defined as BCVA worse than 3/60 in the better-seeing eye.^11^ Severe VI was defined as distance BCVA worse than 6/60 but better than or equal to BCVA 3/60 in the better-seeing eye. Moderate VI was defined as distance BCVA worse than 6/18 but better than or equal to BCVA 6/60 in the better-seeing eye. For better-seeing eye with VI and cataract as the primary cause, the subject was recognized as having ‘VI due to cataract’^11^.

CSC and eCSC were calculated based on the 2022 version of definition.^1, 12^ CSC estimates people with operated cataract (aphakia or pseudophakia) as a proportion of people with operated cataract plus people with cataract and BCVA worse than a specified surgical threshold (also referred to as operable cataract). CSC was calculated using the formula CSC=(x+y)/(x+y+z), where x is the number of individuals with unilateral operated cataract and VI in the other eye, y is the number of individuals with bilateral operated cataract, and z is the number of individuals with VI due to cataracts in at least one eye.

eCSC estimates people with operated cataract attaining a defined level of postoperative presenting visual acuity (i.e., with optical correction, if available) as a proportion of the same denominator. eCSC was calculated using the formula eCSC=(a+b)/(x+y+z), where a is the number of individuals with unilateral operated cataract who have achieved a specified postoperative visual acuity in the operated eye and have VI in the other eye, b is the number of individuals with bilateral operated cataract who have achieved the specified postoperative visual acuity in at least one eye.

Subject flow through the study is shown in Figure 1. In cases where both eyes were operated on, BCVA in the better eye was used to define postoperative visual acuity. Relative quality gap was calculated as (CSC–eCSC)/CSC.^1^

**Figure 1.**
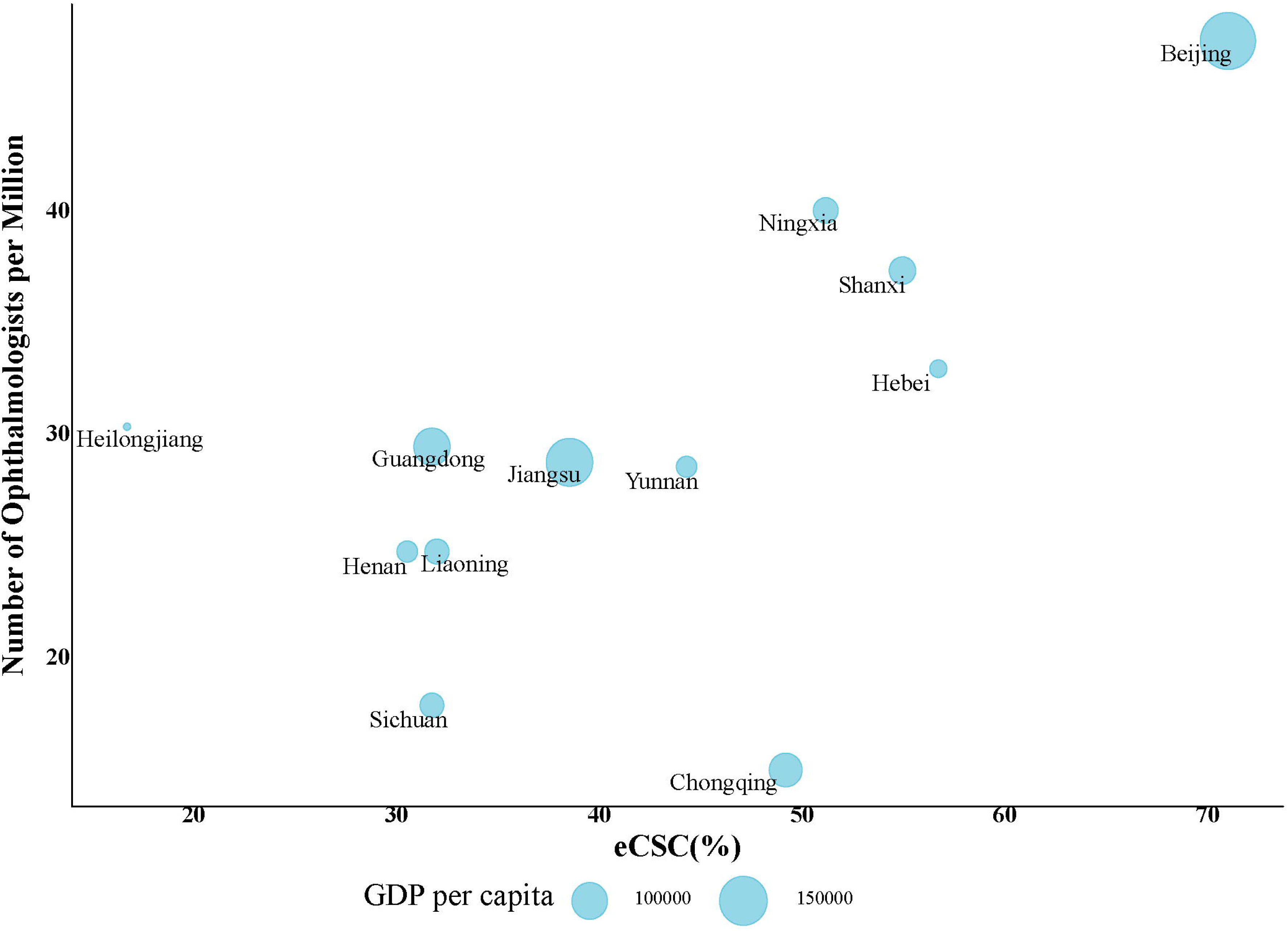
Flowchart depicting categorization of study participants based on cataract surgery status, visual acuity and cause of vision impairment. The variables in the figure (x1, x2, y, z, a, b) refer to specific categories of subjects within the study and are part of formulas to calculate the cataract surgical coverage (CSC) and effective cataract surgical coverage (eCSC). x comprises individuals with one eye with vision impairment(VI, BCVA≥6/18) and the contralateral eye with cataract surgery. x is composed of x1—individuals with unilateral operated cataract achieving a postoperative PVA≥6/18 in the operated eye, and x2—those not achieving this threshold. y includes individuals with bilateral operated cataract, irrespective of the visual acuity outcome. z represents those with VI in both eyes with cataract as the main cause of vision impairment in one or both eyes. a involves individuals with unilateral operated cataract who achieve 6/18 of postoperative presenting visual acuity in the operated eye with VI in the other eye, and b consists of individuals with bilateral operated cataract who has PVA≥6/18 in at least one eye. CSC=(x+y)/(x+y+z). eCSC=(a+b)/(x+y+z). PVA, presenting visual acuity. BCVA, best-corrected visual acuity.

### Statistical analysis

CSC and eCSC are reported using 6/18 as the threshold unless stated otherwise. All estimates were stratified based on the age-sex structure of the population aged 50 and older in the survey area.^13^ Age- and sex-adjusted estimates (age-adjusted only for sex-disaggregated estimates) were calculated from individual participant level data. This involved using the numerators and denominators for VI, CSC and eCSC in age-sex strata (male and female in 10-year bracket). These values were then multiplied by an adjustment factor per stratum (number examined in the sample divided by the number in the population). Finally, adjusted coverage was calculated from the summed outputs. The calculation of the 95% confidence interval (CI) was based on the assumption that the data followed a normal distribution within each specific region; national estimates of 95% CI were calculated using the svymean and confint functions of survey package.^14^

Regional disparity in CSC and eCSC was analyzed using Pearson correlation analysis that included the following factors: a) healthcare resource density index,^15^ e.g., number of registered ophthalmologists (in total/with senior title/with bachelor degree) per million,^16, 17^ b) level of medical resources allocation, e.g., number of registered doctors, health expenditure per capita, top-tier hospitals per capita, ^13^ c) traditional socio-economic indices, e.g., GDP per capita, education coverage, socio-demographic index ^18^ and birth rate.^13^

Logistic regression models were employed to evaluate the association between CSC, eCSC and gender, adjusting for age. Analyses were conducted both comprehensively and separately for each of the 12 provinces/municipalities using the following model:

ecsc∼age+gender

The models were fitted using the glm function, specifying the binomial family. Participants with missing data were excluded. All statistical analyses were conducted using R software (version 4.2.3).

## Results

The current study included 43,045 participants. The number of participants in each of the 12 provinces/municipalities ranged from 1,494 to 6,885 (eFigure1). The age-sex-standardized prevalence of moderate to severe VI and blindness was 4.4% (95% CI: 4.2-4.6) (eTable 2). Cataracts accounted for 39.6% of blindness, 51.0% of severe VI and 54.6% of moderate VI. Among Chinese population aged 50 years and above, which includes approximately 21.6 million individuals suffering from moderate to severe VI and blindness, cataracts were the primary contributing factor in approximately 11.4 million cases.

In our analysis, CSC and eCSC were evaluated at three distinct visual acuity thresholds. At the 6/60 visual acuity threshold, the weighted CSC and eCSC were 90.9% and 84.9%, respectively; the relative quality gap was 6.6% (Table 1). At the 6/18 threshold, the weighted CSC and eCSC were 57.2% and 38.7%, respectively; the relative quality gap was 32.3%. At the 6/12 threshold, the weighted CSC and eCSC were 51.3% and 29.3%, respectively; the relative quality gap was 42.9%.

**Table 1.** Estimates of Regional Effective Cataract Surgical Coverage (eCSC), Cataract Surgical Coverage (CSC), and Relative Quality Gaps at 6/12, 6/18, and 6/60 Visual Acuity Thresholds Across China.

| Province | Adjusted<sup>a</sup><br>CSC(%) | [lower<br>95%CI,upper<br>95%CI] | Adjusted<sup>a</sup><br>ECSC<6/18(%) | [lower<br>95%CI,upper<br>95%CI] | Adjusted<sup>a</sup><br>Gap(%) |
| --- | --- | --- | --- | --- | --- |
| **6/60** |  |  |  |  |  |
| **Weighted** | **90.9** | **86.5-95.2** | **84.9** | **80.6-89.2** | **6.6** |
| Beijing | 99.7 | [99.5,100] | 85.2 | [83.4,87] | 14.5 |
| Chongqing | 97.9 | [97.4,98.4] | 92.3 | [91.4,93.2] | 5.7 |
| Guangdong | 90.3 | [89.6,91.1] | 82.4 | [81.3,83.4] | 8.8 |
| Hebei | 97.9 | [97.4,98.3] | 91.1 | [90.2,92] | 6.9 |
| Heilongjiang | 96.7 | [95.6,97.9] | 96.7 | [95.6,97.9] | 0.0 |
| Henan | 80.2 | [78.9,81.5] | 78.5 | [77.2,79.9] | 2.1 |
| Jiangsu | 90.2 | [89.5,90.9] | 84.7 | [83.9,85.6] | 6.1 |
| Liaoning | 79.9 | [78.5,81.3] | 70.1 | [68.5,71.7] | 12.3 |
| Ningxia | 97.1 | [96.5,97.7] | 95.7 | [95,96.4] | 1.5 |
| Shanxi | 99.2 | [98.8,99.5] | 96.2 | [95.4,96.9] | 3.0 |
| Sichuan | 90.1 | [89.3,90.8] | 80.3 | [79.3,81.3] | 10.8 |
| Yunnan | 100.0 | [100,100] | 98.5 | [98,99] | 1.5 |
| **6/18** |  |  |  |  |  |
| **Weighted** | **57.2** | **47.6-66.8** | **38.7** | **31.7-45.6** | **32.3** |
| Beijing | 87.1 | [85.4,88.8] | 71.0 | [68.7,73.3] | 18.5 |
| Chongqing | 61.2 | [59.5,62.8] | 49.2 | [47.5,50.9] | 19.6 |
| Guangdong | 50.2 | [48.9,51.6] | 31.8 | [30.5,33] | 36.8 |
| Hebei | 87.3 | [86.2,88.3] | 56.7 | [55.2,58.3] | 35.0 |
| Heilongjiang | 52.7 | [50.2,55.1] | 16.7 | [14.9,18.6] | 68.2 |
| Henan | 41.2 | [39.6,42.8] | 30.5 | [29,32.1] | 25.9 |
| Jiangsu | 48.7 | [47.5,49.8] | 38.5 | [37.4,39.7] | 20.8 |
| Liaoning | 52.0 | [50.3,53.8] | 32.0 | [30.4,33.6] | 38.5 |
| Ningxia | 69.4 | [67.8,71] | 51.2 | [49.4,52.9] | 26.3 |
| Shanxi | 61.4 | [59.6,63.3] | 55.0 | [53,56.9] | 10.6 |
| Sichuan | 57.1 | [55.9,58.4] | 31.8 | [30.6,33] | 44.4 |
| Yunnan | 60.2 | [58.2,62.2] | 44.3 | [42.3,46.4] | 26.3 |
| **6/12** |  |  |  |  |  |
| **Weighted** | **51.3** | **40.1-61.7** | **29.3** | **21.8-36.7** | **42.9** |
| Beijing | 84.0 | [82.2,85.9] | 55.7 | [53.1,58.2] | 33.8 |
| Chongqing | 57.4 | [55.7,59.1] | 44.1 | [42.4,45.8] | 23.2 |
| Guangdong | 39.7 | [38.4,41] | 17.0 | [16,18] | 57.2 |
| Hebei | 86.0 | [84.9,87] | 48.2 | [46.6,49.7] | 44.0 |
| Heilongjiang | 37.6 | [35.2,40] | 11.7 | [10.1,13.3] | 68.8 |
| Henan | 39.7 | [38.2,41.3] | 22.6 | [21.2,24] | 43.2 |
| Jiangsu | 42.8 | [41.6,43.9] | 27.7 | [26.6,28.7] | 35.3 |
| Liaoning | 49.0 | [47.3,50.8] | 26.4 | [24.9,28] | 46.1 |
| Ningxia | 65.4 | [63.7,67.1] | 46.7 | [44.9,48.4] | 28.6 |
| Shanxi | 56.2 | [54.3,58.1] | 48.8 | [46.9,50.8] | 13.1 |
| Sichuan | 47.6 | [46.3,48.9] | 20.8 | [19.7,21.8] | 56.4 |
| Yunnan | 54.1 | [52.1,56.1] | 37.1 | [35.1,39.1] | 31.4 |

<sup>a</sup>Weighted prevalence and 95% CI were calculated using the svymean and confint functions of survey package

The underlying causes for the relative quality gap are shown in eTable3, including other ocular diseases(40.5%), PCO (15.6%) and refractive errors (9.0%). Both relative quality gap and refractive error was associated with older age (p<0.001 and p = 0.006, respectively).

Comparison to historical data in and 2014^19^ demonstrated significant improvements in both CSC and eCSC. The weighted change per decade was 38.9% and 26.1% for CSC and eCSC, respectively(Table 2).

**Table 2.** Comparative Trends in Cataract Surgical Coverage (CSC) and Effective Cataract Surgical Coverage (eCSC) at the Visual Acuity Threshold of 6/18 from 2014 to 2022.

|  | CSC |  |  | eCSC |  |  |
| --- | --- | --- | --- | --- | --- | --- |
|  | 2014<sup>a</sup><br>Survey(%) | 2022<br>Survey(%) | Weighted<br>change per<br>decade<sup>b</sup>(%) | 2014<sup>a</sup><br>Survey(%) | 2022<br>Survey(%) | Weighted change<br>per decade<sup>b</sup>(%) |
| Province |  |  |  |  |  |  |
| Beijing | 48.2 | 87.1 | 48.6 | 37.9 | 71.0 | 41.4 |
| Jiangsu | 29.7 | 48.7 | 23.7 | 20.4 | 38.5 | 22.6 |
| Guangdong | 29.7 | 50.2 | 25.7 | 20.5 | 31.8 | 14.1 |
| Heilongjiang | 22.9 | 52.7 | 37.2 | 14.5 | 16.7 | 2.8 |
| Hebei | 31.5 | 87.3 | 69.7 | 21.2 | 56.7 | 44.4 |
| Chongqing | 32.0 | 61.2 | 36.5 | 25.1 | 49.2 | 30.2 |
| Yunnan | 25.1 | 60.2 | 43.8 | 15.7 | 44.3 | 35.8 |
| Ningxia | 31.3 | 69.4 | 47.6 | 22.8 | 51.2 | 35.5 |
| Weighted overall<br>rate | 30.3 | 61.4 | **38.9** | 21.0 | 41.9 | **26.1** |

<sup>a</sup>Statistics of each province in 2014 national survey were not adjusted for age and sex.
<sup>b</sup>Weighted overall rate based on population stratum of each province. Decadal change rate refer to the 2012(estimated)-2022 change rate.

This study revealed significant regional disparity(Figure 2, Table 1). After adjustment for age and sex, the highest CSC was from Hebei at 87.3% (95% CI 86.2-88.3) and the lowest was from Henan at 41.2% (95% CI 39.6-42.8). The highest eCSC estimate was from Beijing at 71.0% (95% CI 68.7-73.3) and the lowest was from Heilongjiang at 16.7% (95% CI 14.9-18.6). The smallest relative quality gap was in Shanxi at 10.6% (CSC 61.4%, eCSC 55.0%), and the largest relative quality gap was in Heilongjiang at 68.2% (CSC 52.7%, eCSC16.7%).

**Figure 2.**
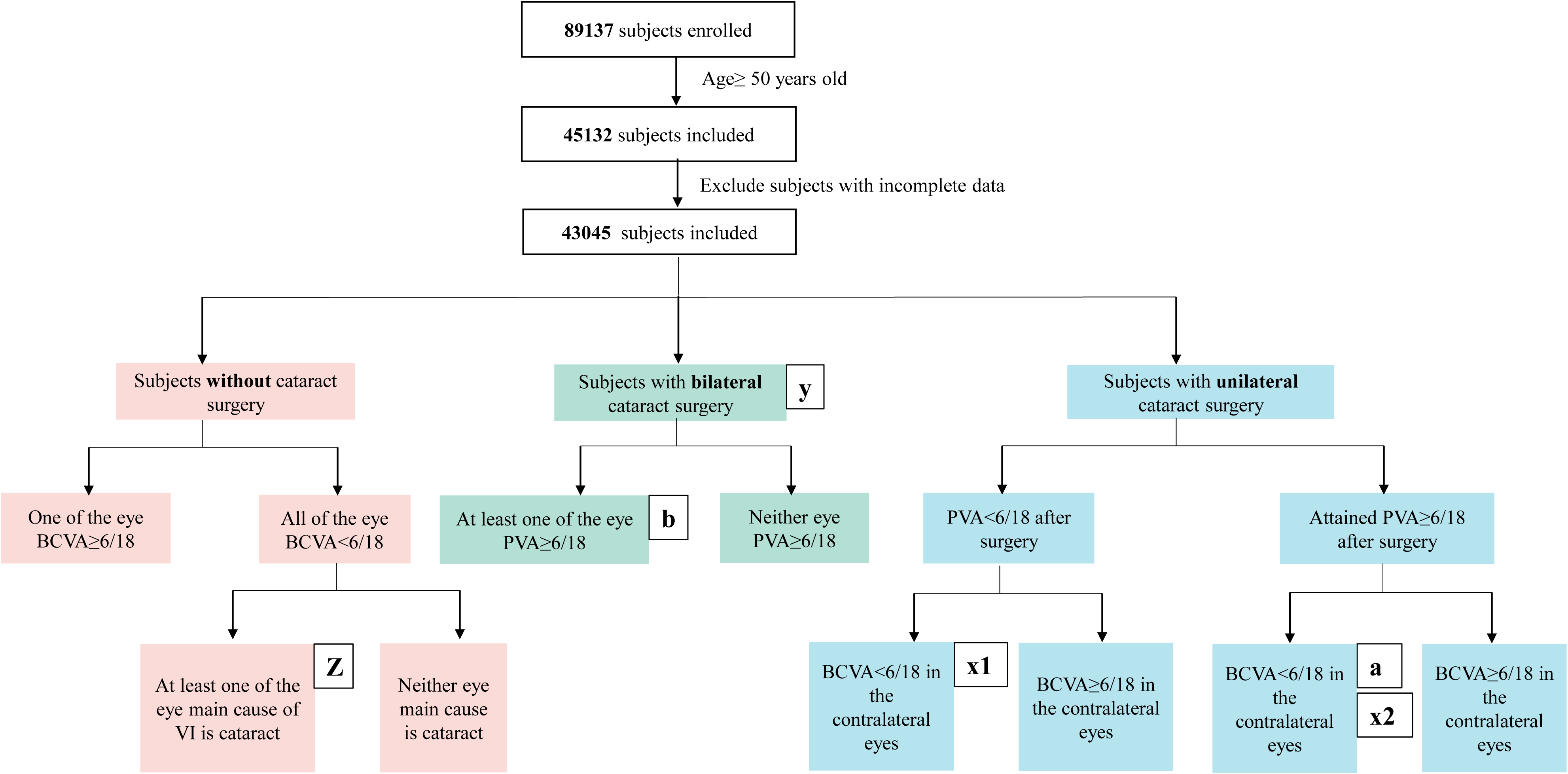
Regional disparity of CSC and eCSC values across different thresholds in China. The thresholds include 6/60, 6/18, and 6/12, which indicate levels of visual acuity required for a ’good’ surgical outcome. CSC, cataract surgical coverage. eCSC, effective cataract surgical coverage.

In Pearson correlation analysis, CSC and eCSC were primarily correlated with the level of medical resources allocation(eTable4 and Figure 3). Notably, there was a strong correlation between eCSC and the availability of doctors (represented by registered doctors per thousand, r=0.64, p=0.026) and ophthalmologists (represented by registered ophthalmologists per million, r=0.58, p=0.047). eCSC also correlated with GDP per capita (r=0.50, p=0.096) and healthcare expenditure per capita (r=0.52, p=0.08). Conversely, CSC and eCSC were irrelevant to indices representing high-quality resources such as the number of top-tier hospitals per person and the presence of senior-titled or highly certified ophthalmologists per million (all p>0.1).

**Figure 3.**
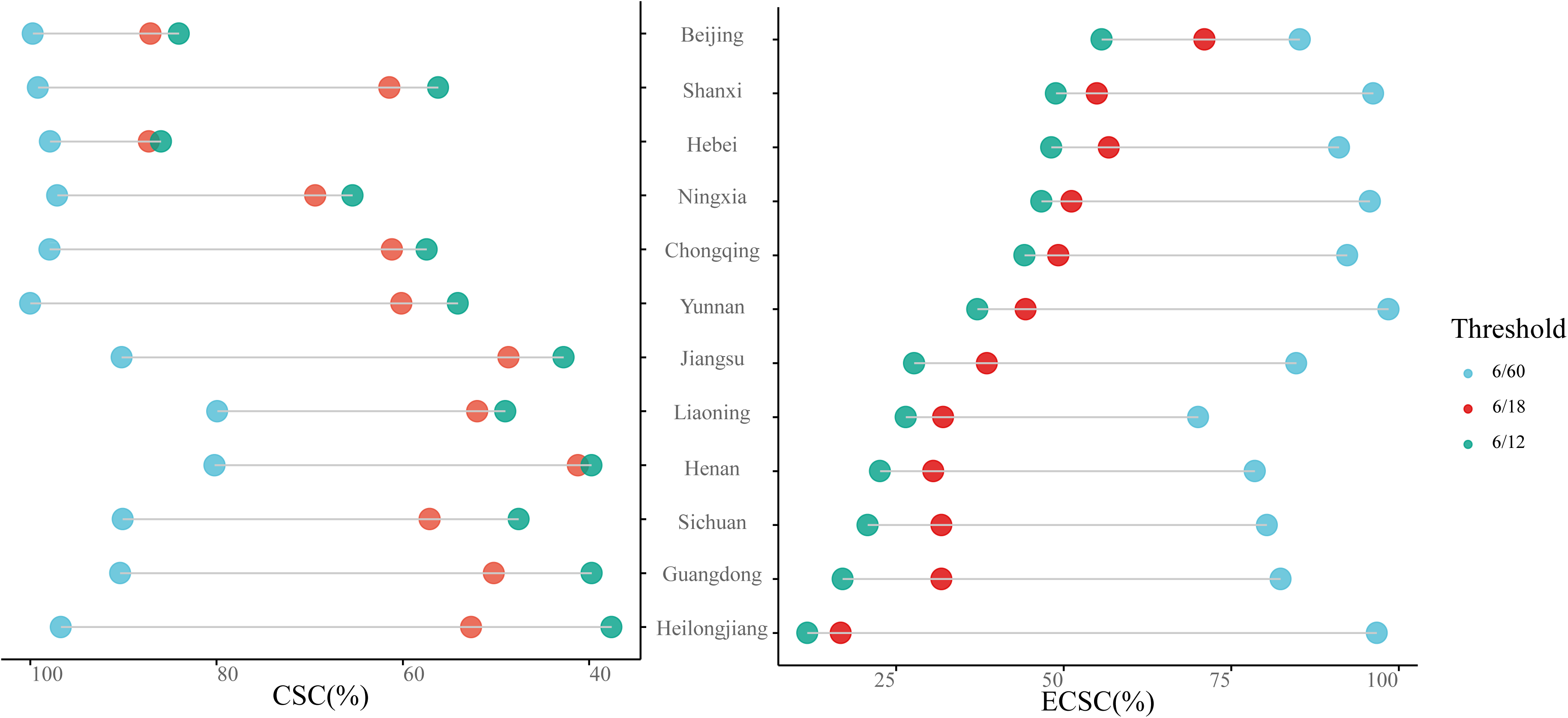
eCSC was primarily correlated with the level of medical resources allocation and was less correlated with the GDP per capita. This graph plots the number of ophthalmologists per million population against eCSC for various provinces in China. Bubble sizes represent the GDP per capita of each province, with larger bubbles indicating higher GDP per capita. eCSC, effective cataract surgical coverage.

The overall analysis demonstrated a statistically non-significant trend indicating lower CSC (OR: 0.90, 95% CI: 0.78-1.04) and eCSC (OR: 0.88, 95% CI: 0.74-1.04) in women. Furthermore, province-specific logistic regression analyses did not reveal significant differences in CSC or eCSC, except for female was inferior in CSC in one place(OR: 0.60, 95% CI: 0.41-0.88) (eTable 5).

## Discussion

This study provided national estimates of CSC and eCSC in 2021-2022 period in China as well as pointed out the significant regional disparity. The availability of medical resources has proven to be a crucial element in enhancing CSC and eCSC rates, serving as a key predictor of access to quality cataract surgery. We also identified subjects with moderate vision impairment with less likelihood to receive surgical coverage. Most importantly, the definition of eCSC should be revisited to avoid bias from concurrent ocular diseases.

The prevalence of blindness and VI has shown a marked reduction in China over recent years. In the 2014 national survey, the blindness rate was recorded at 6.2%, with a VI rate of 2.4%^19^.In contrast, our current study indicates a decline to 3.8% for blindness and 0.6% for VI. Similarly, in 2019,^20^ VI and blindness were estimated to affect 48.4 million Chinese individuals aged 50 and above, respectively. On the contrary, our current estimates suggest that 21.7 million by VI and blindness within this demographic. This significant reduction in the number of people experiencing vision loss may be partly attributable to a lower response rate in our current study (76.2%). Nonetheless, it also likely reflects substantial progress in mitigating reversible vision impairment across China.

Comparison to historical data(5,6) in China demonstrated significant improvements in both CSC and eCSC. Over the past decade, the CSC and eCSC have increased by 38.9% and 26.1%. The improvement in CSC and eCSC over time is encouraging as the increase in eCSC surpasses the decadal change rate available in the world(Nepal, Bhutan and Mexico)^1^. Despite a recent report from rural Guangdong^21^ indicating no progress or even a decline in CSC (2014: 41.7%; 2020: 40.6%) and eCSC (2014: 32.6%; 2020: 26.6%), we believe this finding reflects regional disparity.

Similar to previous studies in other regions,^20, 22, 23^ we observed significant regional disparity in this study. Notably, eCSC correlated significantly with GDP and healthcare expenditure per capita. These findings are consistent with previous studies,^24^ and calls for major effort in allocating more resources to less developed areas. National and provincial programs that prioritize cataract surgery availability for underprivileged groups in China, particularly the Cataract Surgery Project for Millions of Poor Patients,^25^ have been instrumental in reducing cataract-related blindness, particularly in less developed regions. Also, increasing medical resources, especially improving ophthalmological infrastructure is of paramount importance in improving access to cataract surgery.^26–29^ In this study, the availability of ophthalmologists^30^ or physicians strongly correlated with eCSC, especially in low economic regions (e.g., Hebei and Ningxia). Such a finding indicates continuing efforts are needed.

Though higher than median eCSC in the Africa(13.9%),Western-Pacific(21.0%), Americas(29.2%), Eastern Mediterranean(34.9%) and European region(37.7%), the eCSC rate at the 6/18 threshold (38.7%) in China is lower than the median number in South-East Asia Region(40.4%)^1^. Similarly, relative quality gap(32.3%) is higher than median number of most regions except Americas(30.4%) and South-East Asia(27.6%). This emphasizes an urgent need for targeted interventions and a more strategic allocation of resources to improve cataract surgery quality and access.

The substantially lower CSC and eCSC and wider relative quality gap at 6/18 and 6/12 vs 6/60 in this study suggest that many of those with moderate VI(bilateral VI in the 6/60 to 6/18 range) are not receiving surgical operation. Since opacity of the lens interfere with early diagnosis of retinal diseases and decrease quality of life, we believe that policy makers should focus more on those with moderate VI.

The results of the current study highlight the need for more precise preoperative assessment of intraocular lens power, along with the provision of corrective glasses to address postoperative refractive errors. Additionally, improved postoperative follow-up is essential for PCO, which accounted for 24.9% of the relative quality gap. PCO can be easily managed but previous studies have highlighted a notable lack of awareness regarding the treatment of PCO.^31^ Thus, it is crucial to disseminate the information that PCO as long-term complications of cataract surgery are possible and can be easily managed.

Approximately half of the patients dissatisfied with their visual outcomes post-surgery had other ocular conditions (e.g., retinal diseases). Those patients may achieve satisfactory results immediately after cataract surgery but subsequently develop retinal diseases, hence is retinal diseases rather than quality of surgery results in VI. Since eCSC was proposed to pinpoint the quality of surgery, we proposed participants with comorbidities should be excluded in further study.

The strength of the current study lies in the relatively large sample size across representative regions in China. Also, ophthalmic examination was conducted by either ophthalmologists or trained nurses, thus ensuring data accuracy.

The current study has several key limitations. First, the reasons for not getting cataract surgery were not investigated. Second, we did not collect systemic conditions (e.g., diabetes) that may contribute to unsatisfactory postoperative visual outcome. Third, the response rate of current study is 76.2%; accordingly, blindness may be under-estimated. Fourth, the preoperative visual acuity of persons having had surgery before baseline was unknown.

In summary, both CSC and eCSC improved substantially during the past decade in China based on comparison to historical data. However, significant quality gap existed. Also, there was considerable regional disparity. These results call for continuing efforts in improving eCSC, and particularly programs that targeted relatively less developed regions.

## Contributors

JL and KC had full access to all the data in the study and take responsibility for the integrity of the data and accuracy of the data analysis. ZJ and NW conceived and designed the study. JX and ZJ were responsible for fundus diagnosis. XY, SJ, QZ, AH, MZ, QL, HZ, WH, XL, ZS, BS, WZ, XZ, ZH, ZW, HZ, WH helped conduct the study. All authors approved the final version of the report.

## Decalration of competing interests

We declare that there are no competing interests.

## Data sharing

Our statistical code is available on request to the corresponding author.

## Data Availability

All data produced in the present study are available upon reasonable request to the authors

## Acknowledgements

We sincerely thank all the participants and healthcare workers that involved in current study.

